# 4th Dose COVID mRNA Vaccines’ Immunogenicity & Efficacy Against Omicron VOC

**DOI:** 10.1101/2022.02.15.22270948

**Authors:** Gili Regev-Yochay, Tal Gonen, Mayan Gilboa, Michal Mandelboim, Victoria Indenbaum, Sharon Amit, Lilac Meltzer, Keren Asraf, Carmit Cohen, Ronen Fluss, Asaf Biber, Ital Nemet, Limor Kliker, Gili Joseph, Ram Doolman, Ella Mendelson, Laurence S. Freedman, Dror Harats, Yitshak Kreiss, Yaniv Lustig

## Abstract

**BACKGROUND:** Following the emergence of the Omicron variant of concern, we investigated immunogenicity, efficacy and safety of BNT162b2 or mRNA1273 fourth dose in an open-label, clinical intervention trial.

**METHODS:** Primary end-points were safety and immunogenicity and secondary end-points were vaccine efficacy in preventing SARS-CoV-2 infections and COVID-19 symptomatic disease. The two intervention arms were compared to a matched control group. Eligible participants were healthcare-workers (HCW) vaccinated with three BNT162b2 doses, and whose IgG antibody levels were ≤700 BAU (40-percentile). IgG and neutralizing titers, direct neutralization of live VOCs, and T-cell activation were assessed. All participants were actively screened for SARS-CoV-2 infections on a weekly basis.

**RESULTS:** Of 1050 eligible HCW, 154 and 120 were enrolled to receive BNT162b2 and mRNA1273, respectively, and compared to 426 age-matched controls. Recipients of both vaccine types had a ∼9-10-fold increase in IgG and neutralizing titers within 2 weeks of vaccination and an 8-fold increase in live Omicron VOC neutralization, restoring titers to those measured after the third vaccine dose. Breakthrough infections were common, mostly very mild, yet, with high viral loads. Vaccine efficacy against infection was 30% (95%CI:-9% to 55%) and 11% (95%CI:-43% to +43%) for BNT162b2 and mRNA1273, respectively. Local and systemic adverse reactions were reported in 80% and 40%, respectively.

**CONCLUSIONS:** The fourth COVID-19 mRNA dose restores antibody titers to peak post-third dose titers. Low efficacy in preventing mild or asymptomatic Omicron infections and the infectious potential of breakthrough cases raise the urgency of next generation vaccine development.

**Trial registration number:** clicaltrials.gov: NCT05231005, NCT05230953

---

Since December 2019, the SARS-CoV-2 pandemic has rapidly spread globally. One year later, vaccine rolled-out began, concurrently with a third pandemic surge of the Alpha variant of concern (VOC).

The BNT162b2 vaccine was rolled out in Israel, on December 19, 2020. Following the first two doses, high vaccine effectiveness was reported by real-world observational studies. Initial induction in immunogenicity^1^ followed by reduction in infections, disease, hospitalization and death were reported^2–4^, as well as reduction in infectivity among breakthrough cases^5^ By April 3, 2021, over 90% of Israeli adult population aged ±60 was vaccinated with two doses (with a 21 day interval between doses)^6^.

Yet, with time, waning of this protective effect was reported, both in immunogenicity as well as in vaccine effectiveness against infections and hospitalizations^7,8^ resulting in a fourth pandemic surge in Israel dominated by the Delta VOC. These led the Israeli ministry of health (MOH) to rollout a third, BNT162b2 booster dose on July 29, 2021, to adults 60 years and older. Within a few weeks, the campaign expanded to the whole population, in whom at least 5 months have passed from the second dose.

The third dose was shown to be effective in mounting a significant humoral and cellular immune response^9,10^, leading to protection from infection and disease^11^. Many countries later followed the Israeli policy of a third booster dose as part of the vaccine schedule. Yet, within four months after approval of the third dose, on Nov. 29, 2021, the Omicron VOC was first reported in Israel.

The Omicron VOC has rapidly emerged worldwide, with extremely high transmission rates,^12–14^. Although the Omicron VOC, appeared to be less virulent than previous VOCs, its high transmissibility lead to dramatic surges of cases globally and threatened to overwhelm healthcare systems, to a state of collapse. This raised the question of the need for a fourth SARS-CoV-2 vaccine dose. Yet, a modified-Omicron vaccine has not yet been developed. Data on immunogenicity, safety and effectiveness of a fourth dose are therefore urgently needed.

Here, we report findings from an ongoing clinical trial evaluating the safety, immunogenicity and efficacy of a fourth dose of two mRNA vaccines administered after three BNT162b2 doses, in preventing SARS-CoV-2 infections.

## METHODS

### Trial Objectives, Participants and Oversight

This open-label, nonrandomized, clinical trial was performed to assess the safety and immunogenicity of a fourth dose of either BNT162b2, or mRNA1273 COVID-19 vaccine. Eligible participants were persons 18 years of age or older, with no known history of SARS-CoV-2 infection, who received the third dose of the full BNT162b2 schedule, at least 4 months earlier, and who were enrolled to the Sheba HCW COVID-19 Cohort study^8^, and thus with a known immune response history to the previous doses. Full eligibility criteria are available in the supplementary protocol. In order to enroll persons at expected higher risk of infection, the trial population was selected among participants of the serology study cohort who had IgG titers below the 40% percentile at that time, a titer which was 700 BAU or lower (**Figure 1**).

**Figure 1:**
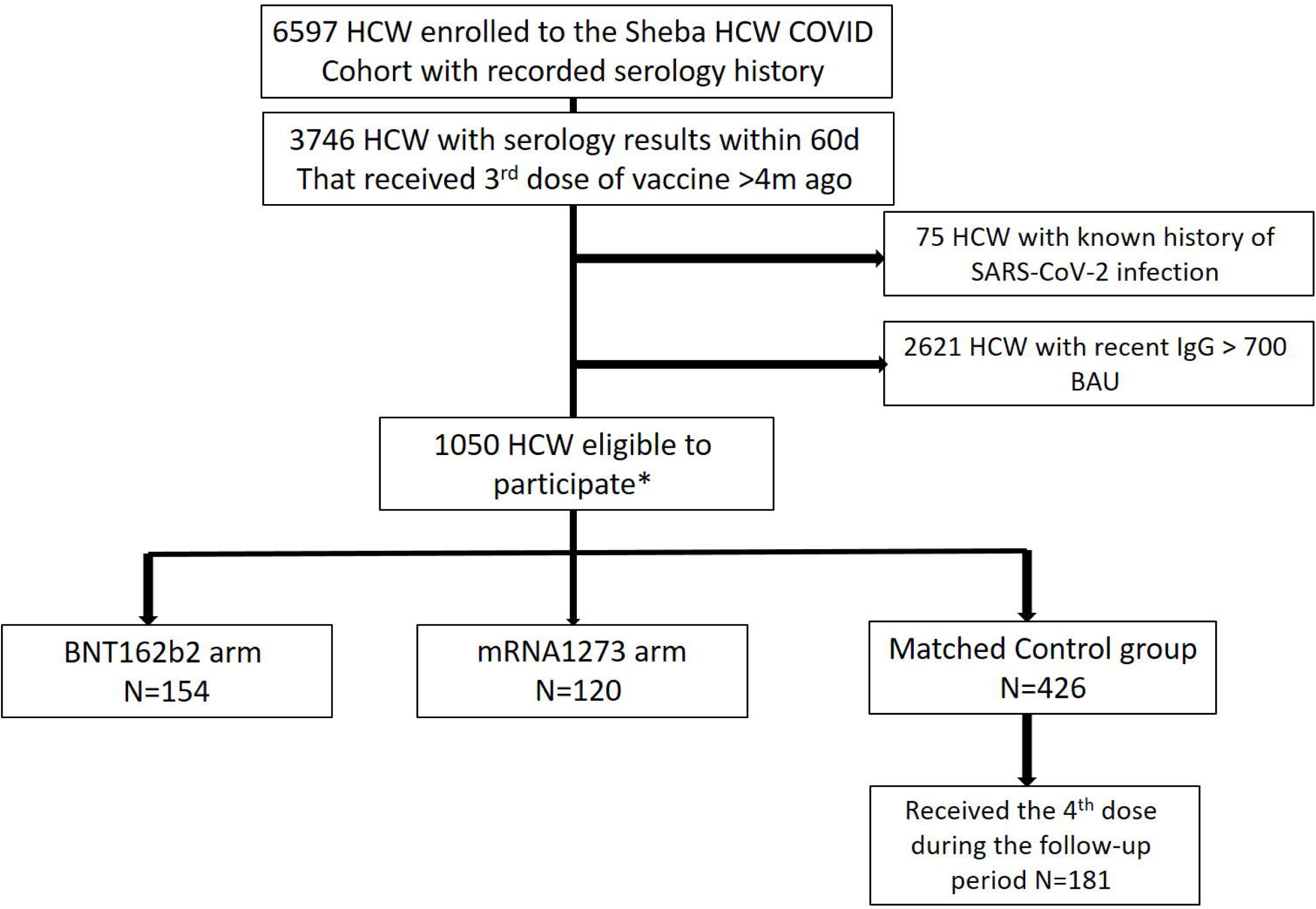
Study source cohort, screening and participant eligibility and enrollment

Participants in the first arm were enrolled to receive a fourth dose of 30µg BNT162b2 on Dec 27-28, 2021. One week later, on Jan 5-6, 2022, addition of the second arm was approved and additional participants were enrolled to receive 50µg mRNA1273 as a fourth dose. An age matched control group was selected from all HCW of the Sheba HCW COVID-19 Cohort who were eligible to be enrolled to one of the intervention arms; i.e., those who received three BNT162b2 doses, at least four months earlier, were recruited to the Sheba HCW COVID Cohort, with a titer of 700 BAU or lower, but did not receive a fourth dose). Two controls were matched by age to each trial participant. For detail, see the Supplementary protocol and Supplementary Methods S1, S2.

The ongoing trial is being conducted in accordance with the International Council for Harmonization of Technical Requirements for Pharmaceuticals for Human Use, Good Clinical Practice guidelines, and applicable government regulations. The national and the institutional review board approved the protocol and the consent forms. All participants provided written informed consent before enrollment. Safety is reviewed by an independent data and safety monitoring board on a weekly basis. This is an independent study, not sponsored or funded by any commercial company. All trial vaccines were acquired through the government procurement process.

### Trial Procedures and Study Period

Enrollment to the two arms was time dependent, those enrolled by December 28 joined the BNT162b2 arm and those enrolled later, until January 6, joined the mRNA1273 arm. Maximal enrollment into each arm was 175 subjects. After informed consent was obtained, participants underwent screening and medical and vaccine history were collected (Supplementary **Table S1**). Blood samples for immunogenicity assessments were collected, a nasopharyngeal swab for SARS-CoV-2 PCR was obtained and the designated vaccine dose was administered; either 30µg of BNT162b2, for those enrolled on December 27-28, 2021, or 50µg of mRNA1273 for those enrolled on January 5-6, 2022.

Follow up visits took place on days 7, 14 and 21 and included safety assessment, symptom screen, SARS-CoV-2 PCR nasopharyngeal swab and blood for immunogenicity tests as detailed below. A final assessment of symptoms and SARS-CoV-2 testing (either PCR or rapid Ag tests) was performed on day 30, by telephone, electronic questionnaires and laboratory SARS-CoV-2 testing database.

### Safety

Safety assessments included monitoring of immediate and late solicited and unsolicited adverse events. Immediate adverse events and allergic reactions were monitored for 30 minutes after vaccine administration, by study physicians or nurses. Solicited adverse event data were collected by an electronic questionnaire distributed on day 5 after vaccination and on a weekly basis for three consecutive weeks (See Supplementary **Table S2**). Any participant who did not submit the electronic questionnaire, was directly contacted by research coordinators via telephone calls and missing data was retrieved. Solicited AE included any local reactions as well as systemic reactions, including fever, fatigue, myalgia, headache, lymphadenopathy and other systemic reactions. Severity and duration of any symptom were reported. Participants were instructed to report any unsolicited AE, any medically attended AE, need for medication, ED visits or hospitalization, within the study period.

### Immunogenicity

The primary end points were the immunogenicity of the fourth dose of both vaccines, and specifically assessment of in-vitro neutralization of omicron VOC. Immune responses tested include: (i) SARS-CoV-2 IgG II Quant (Abbott, IL, USA), (ii) SARS-CoV-2 pseudovirus neutralization assay using a vesicular stomatitis virus (VSV) backbone coated with SARS-CoV-2 spike (S) protein (ref), (iii) Live microneutralization of different strains; Wu-1 as well as Alpha, Delta and Omicron VOCs ^15^. T cell activation was tested by Enzyme-linked immune absorbent spot (Elispot) to measure antigen-specific T cells that secrete Interferon-γ, on two time points; visit 1, before 4^th^ dose administration and visit 3, two weeks later. Additional detail is provided in the Supplementary Methods S3.

### Vaccine efficacy

Secondary end points were the fourth dose vaccine efficacy in preventing infection and symptomatic disease compared to three vaccine doses only. To identify any infection, whether symptomatic or not, nasopharyngeal swabs were obtained on each weekly visit, for SARS-CoV-2 by RT-PCR (Seegene, South Korea). The control group was urged via text message reminders to undergo weekly screening PCR tests regardless of symptoms. In addition, participants in both arms and in the control group, were requested to perform a SARS-CoV-2 test (either RT-PCR or rapid Ag test) in case of any event of exposure to a detected SARS-CoV-2 infected person or if they developed any potential COVID-19 symptom, including fever, sore throat, headache, myalgia, rhinorrhea, cough or loss of smell or taste. Symptoms were assessed on each weekly visit. All PCR SARS-CoV-2 tests conducted in the hospital or in other settings were reported to a central reporting system, and participants were actively inquired about results of home rapid antigen tests (via electronic questionnaires or telephone calls).

Breakthrough cases were defined only from day 8 (in each arm as well as in the control group), to exclude early infections due to exposure before vaccine is effective. All breakthrough cases were assessed by electronic questionnaires or telephone calls to define symptom severity at the end of their infection period.

### Statistical Analysis

#### Safety and Immunogenicity

Safety results and analyses are descriptive, reporting point estimates with 95% confidence levels. To compare adverse events between the two intervention groups, we stratified the adverse events by age. Similarly point estimates of IgG and neutralizing antibody titers were reported as GMT with 95%CI. T cell activity/million cells was reported as mean with 95%CI.

#### Vaccine efficacy

The cumulative incidence of infections and of symptomatic disease of each intervention arm were calculated and compared to its control group, starting from the eighth day after the vaccine was first administered and ending on 30^th^ January 2022. The start of follow-up for the controls was January 3, 2022 for BNT162b2 (eighth day after December 27, 2021), and January 12, 2022 for mRNA1273 (eighth day after January 5, 2022). Controls who received a fourth vaccine dose during the follow-up were censored on the day they received the vaccine. The infection incidence rate ratio (vaccine group versus control) and vaccine efficacy (1 minus the rate ratio) were estimated using a Poisson regression model that adjusted for calendar day and age group (18-45, 46-60, >60y), including persons in the vaccine groups from the eighth day following receipt of the vaccine. In a secondary analysis, rate ratios and vaccine efficacies were estimated for two separate periods following the vaccines: 8-14 days and ≥15 days. Further details are provided in the Supplementary Appendix.

## RESULTS

### Participants

Of a total of 6,597 HCW enrolled to the Sheba HCW COVID Cohort since 2020, 1050 were eligible to participate (Figure 1). Of these, 154 were enrolled to the BNT162b2 arm on December 27-28, 2021. One week later, on January 5-6, 2022, after the second arm was approved, 120 participants were enrolled to the mRNA1273 arm. From the remaining eligible 776 participants, controls were age matched in a 2:1 ratio to participants in each arm. Participants in the BNT162b2 arm were slightly older than those in the mRNA1273 arm (58.9±13.3, vs. 55.1±12.5). Baseline characteristics of the study trial population on enrollment is described in **Table S3**.

On January 3, 2022, the Israeli MOH approved a fourth vaccine to any adult older than 60, and to all HCW. Thus, from the Control group of 426, during the study period 181 received the fourth vaccine out of the study, and after vaccination were censored and contributed no further information to our analyses.

### Fourth dose Safety

No immediate responses were recorded. All 274 participants in both BNT162b2 and mRNA1273 arms replied to the adverse event questionnaire sent on day 5, 7, 14 and 21. No serious adverse events were reported. No hospital admissions were reported. The only unsolicited events reported were ocular redness and pain by two participants who were referred to the ophthalmology emergency room, where they were examined and diagnosed with conjunctivitis, unrelated to vaccination.

Solicited local adverse events (AE) were common, reported by 121 (78.6%, 95%CL: 71.2-84.8%) of BNT162b2 recipients vs. 99 (82.5%, 95%CL 74.5-88.8%) of mRNA1273 recipients. Among BNT162b2 recipients, local AE were more often reported by younger participants (88% 95%CL 80.6-95.3% compared with 69.6%, 95%CL 59.4-79.7% in >60 years of age). This difference was minor among mRNA1273 recipients. Most AE were reported as mild, and resolved within 1.7 days (**Figure 2, Table S4**).

**Figure 2:**
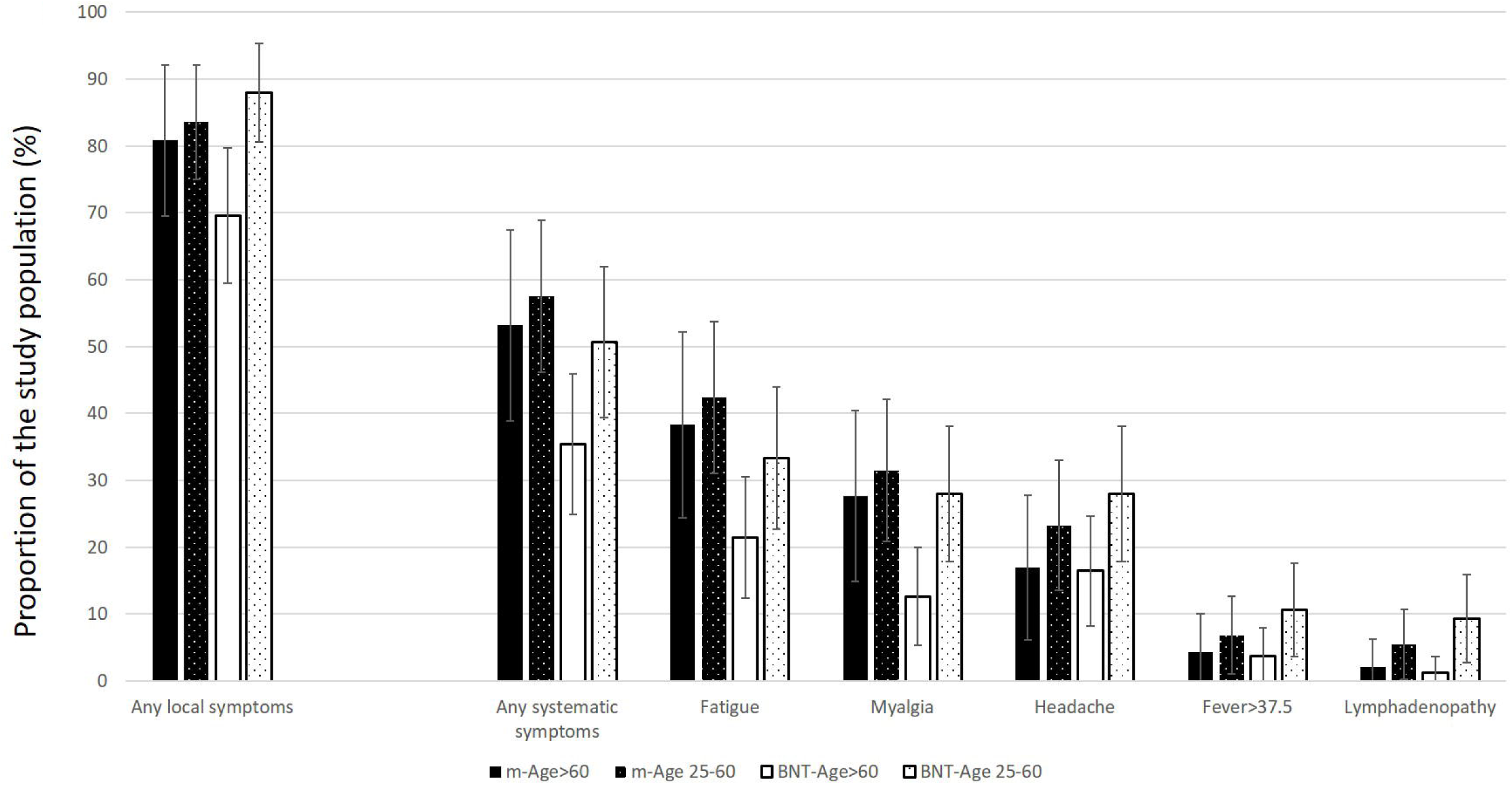
Solicited local and systemic adverse events following the 4th dose of BNT162b2 or mRNA1273. Black denotes recipients of mRNA1273, white denotes recipients of BNT162b2. Dotted denotes the younger population (<60 years of age). The proportion of participants reporting each AE among each age group in the two different vaccine recipients, is presented and its 95%CL.

Solicited systemic adverse events were reported by 42.9% (95%CL 35-50.7%) of BNT162b2 recipients and 55.8% (95%CL 46.9%-64.7%) of mRNA1273 recipients. Younger participants reported systemic AE more commonly than older participants for each of the AEs and in both vaccines, but this effect was small and did not reach statistical significance. Systemic adverse events resolved within 1.3 days ± 2.42. The most common adverse event reported was fatigue (27.3%, 95%CL 20.4-35.0% in BNT162b2 and 40.8%, 95%CL 31.9-49.6% in mRNA1273), followed by myalgia and headache. Fever was relatively uncommon, with only 7.1% (95%CL 3.1-11.2%), reported fever above 37.5°C, and slightly less in mRNA1273 recipients. Fever resolved within 24-36h in either group (**Figure 2, Table S4**).

### Immunogenicity

Five months after receipt of the third dose (pre-dose 4) anti-RBD IgG titers were 6-fold lower than one month after receiving the third dose (peak post-dose-3 titers) in all three groups. Yet, they were 5 fold higher than titers measured five months after the second dose (pre-dose 3). Within 1-3 weeks of administration of either of the vaccines’ fourth dose, anti-RBD IgG titers increased 9-10-fold, to titers slightly higher than those of the first month after the third dose (Table S5 and Figure 3). At the same time, anti-RBD-IgG levels of the control group continued to wane (IgG GMT level of 340, 95%CI: 303-381).

**Figure 3:**
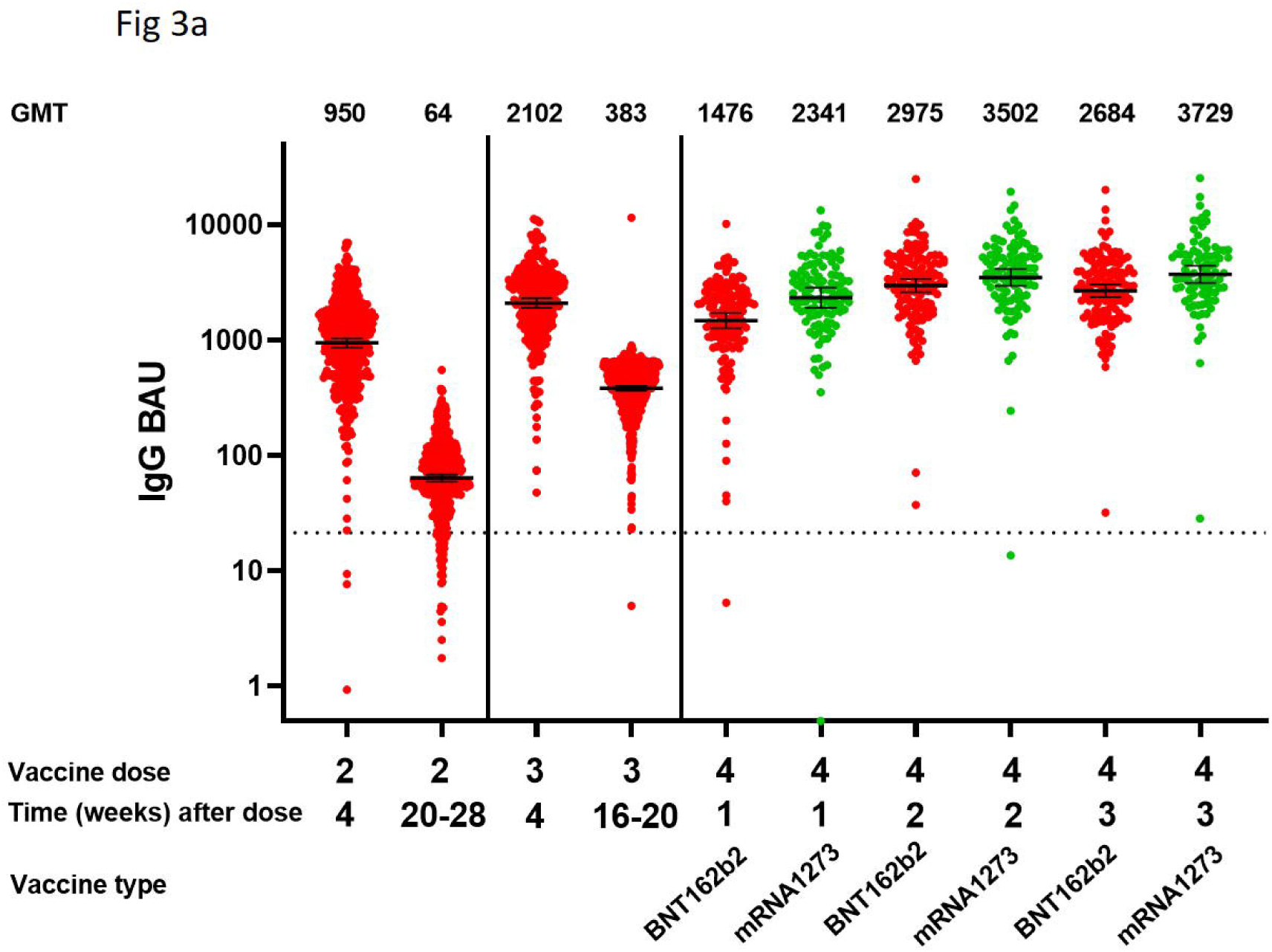
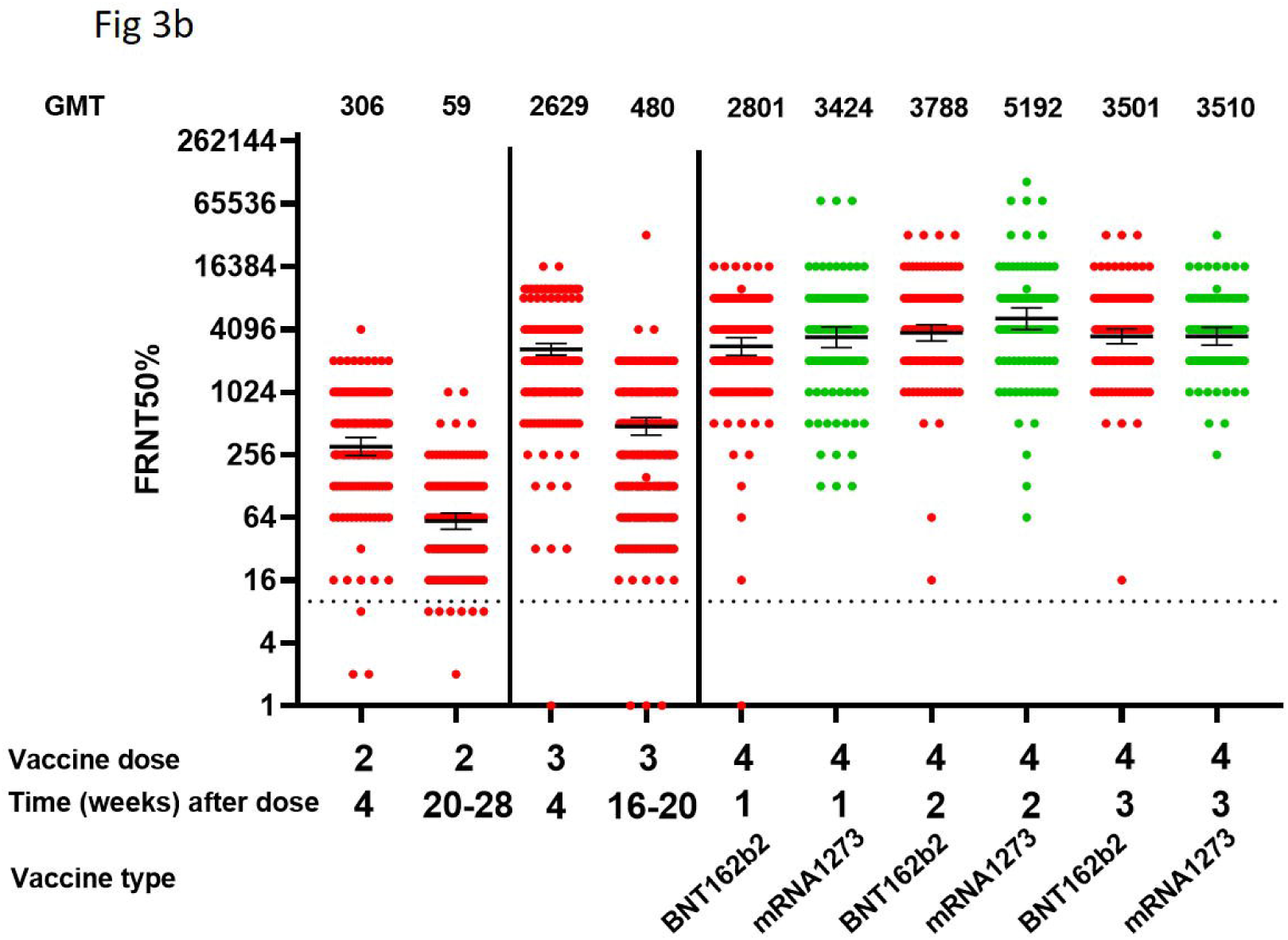
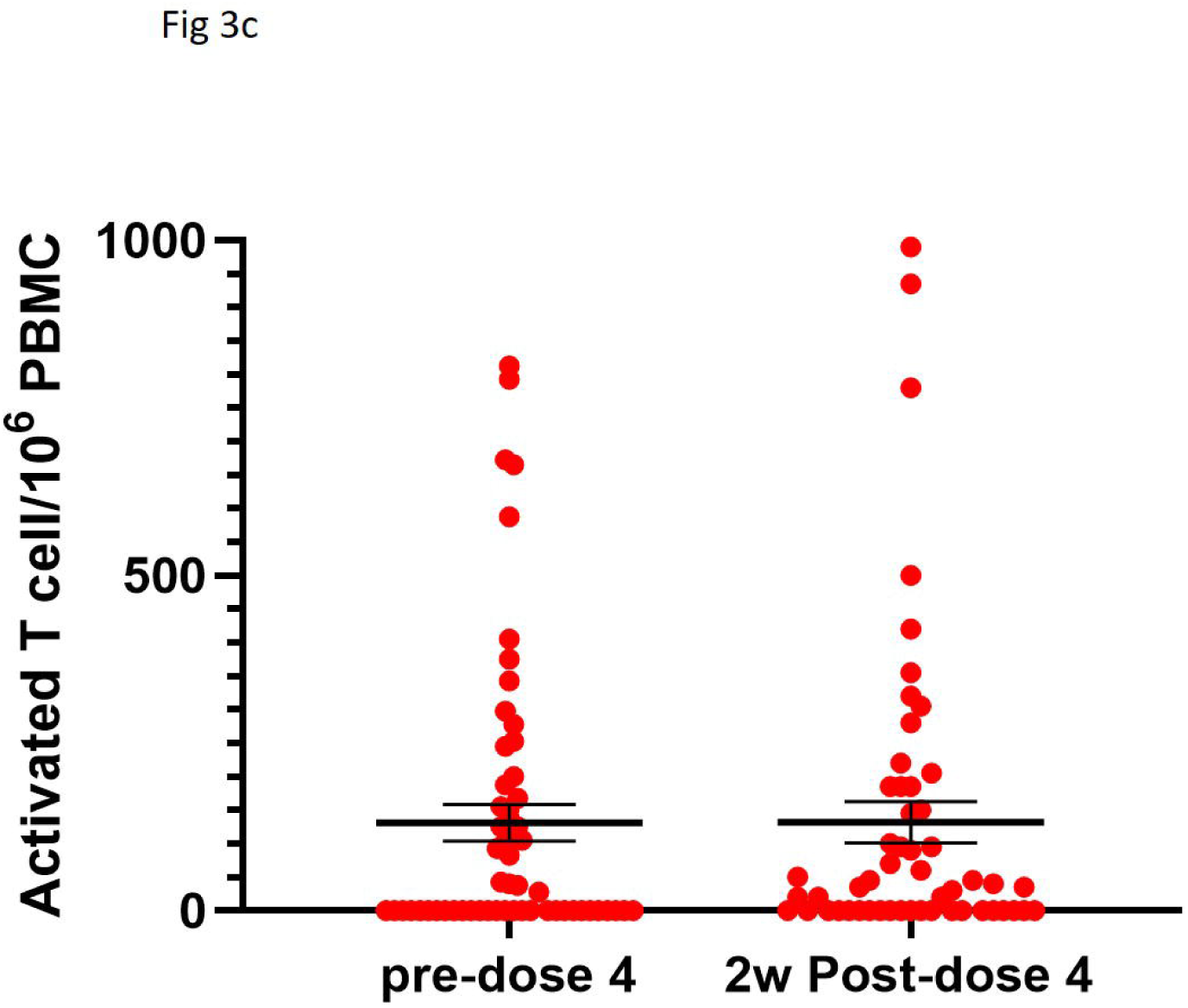
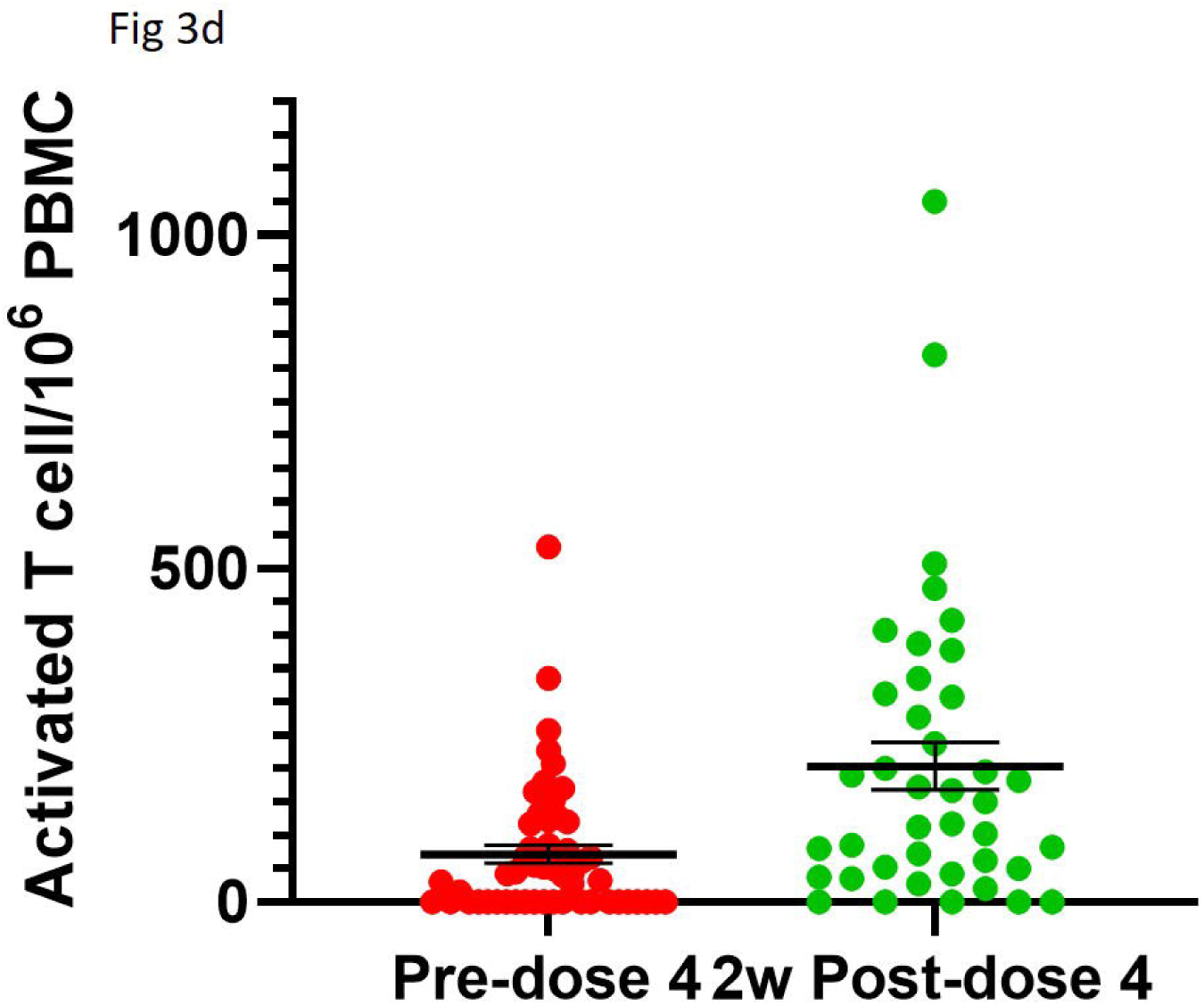
Immune response after each dose a. IgG titers after 3 doses of BNT162b2, and a fourth dose of either BNT162b2 or mRNA1273, b. Neutralizing antibody titers after 3 doses of BNT162b2, and a fourth dose of either BNT162b2 or mRNA1273, c. T cell activation after 4 doses of BNT162b2, d. T cell activation after 3 doses of BNT162b2 and a fourth dose of mRNA1273. Geometric mean titers (GMT) are presented and their 95%CI. Red denotes response among BNT162b2 recipients; Green denotes response among mRNA1273 recipients.

All but two cases of pre-dose-4 neutralizing antibody titers (4-5 months after the third dose) were above the limit of detection, with a GMT of 355 (95%CI 270-467) and 276 (95%CI 210-363) for BNT162b2 and mRNA1273, respectively. At this time point, they were 7-9-fold lower than at their peak after the third dose. Yet, they were 5-6-fold higher than at their pre-dose-3 time point (five months after the second dose). Within two weeks of administration of the fourth dose a peak level was observed, reaching 3788 IU and 5192 IU for BNT162b2 and mRNA1273, respectively (**Table S5, Figures 3c and d)**.

### T cell response

In total, 58 and 56 recipients of BNT162b2 and mRNA1273, respectively, were assessed for T-cell activation on day 1, before receiving the fourth dose. Among BNT162b2 recipients, 53 of the 56 were re-assessed on day 14. The proportion of responders increased from 50% to 60%, yet, the mean number of T cells activated by the spike protein did not change (131±27 to 132±32). Among mRNA1273 recipients, 40 of the 56 were re-assessed on day 14. The proportion of responders increased from 61% to 87% and the average IFNγ activated T cells increased from 72±13 to 203±36 (**Table S5, Figure 3e-3f**).

### Direct neutralization of Omicron compared to other strains

Samples of 25 randomly picked participants in each group were assessed for direct neutralization of Omicron VOC compared to Delta VOC and Wu-1 strain before the fourth dose (4-5 months after the third dose), and 7 and 14 days after. For both vaccines, and at all time-points, neutralization of Omicron was approximately 10-fold less than that of Wu-1 and 4-7 folds lower than Delta. BNT162b2 recipients demonstrated increased neutralization of Omicron VOC by 8.5-fold, one week after administration of the fourth dose, with an additional non-significant increase during the second week, reaching a 10.7-fold increase by day 14. mRNA1273 recipients demonstrated a 7-fold increase within one week that did not increase much further by day 14 (total 7.2-fold increase). Among BNT162b2 recipients Delta neutralization increased by 11-fold within one week, with no further increase observed at 14 days. While a 10.1-fold increase was observed for mRNA1273 recipients within 1 week, that further increased to 15.6-fold by day 14 (Figure 4a and 4b).

**Figure 4:**
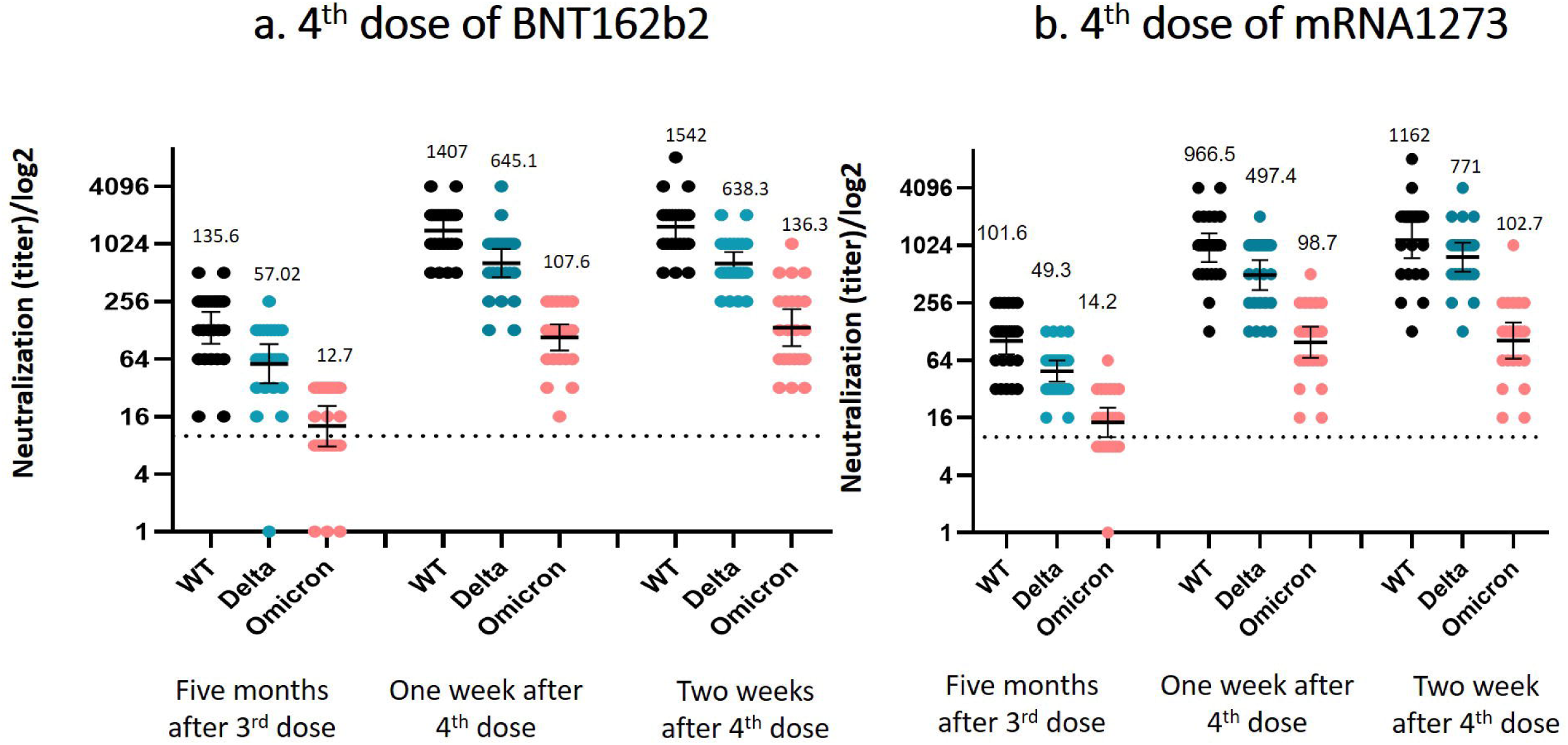
Live virus neutralization efficiency against different strains on the different time points; Black denotes the wild type (Wu-1), blue represents neutralization of the Delta VOC, red represents the neutralization of Omicron. GMT with their 95%CI are depicted.

**Figure 5:**
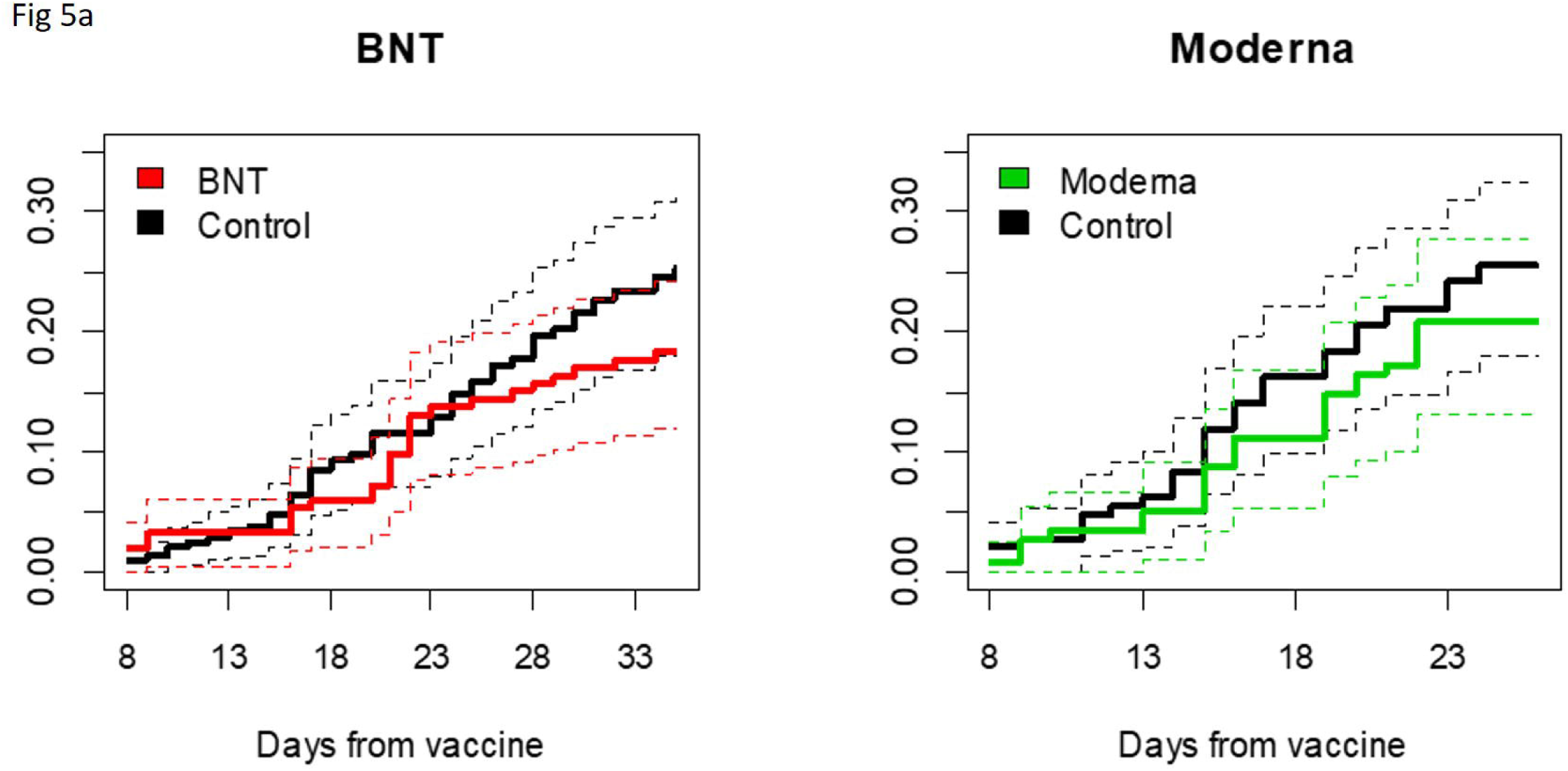
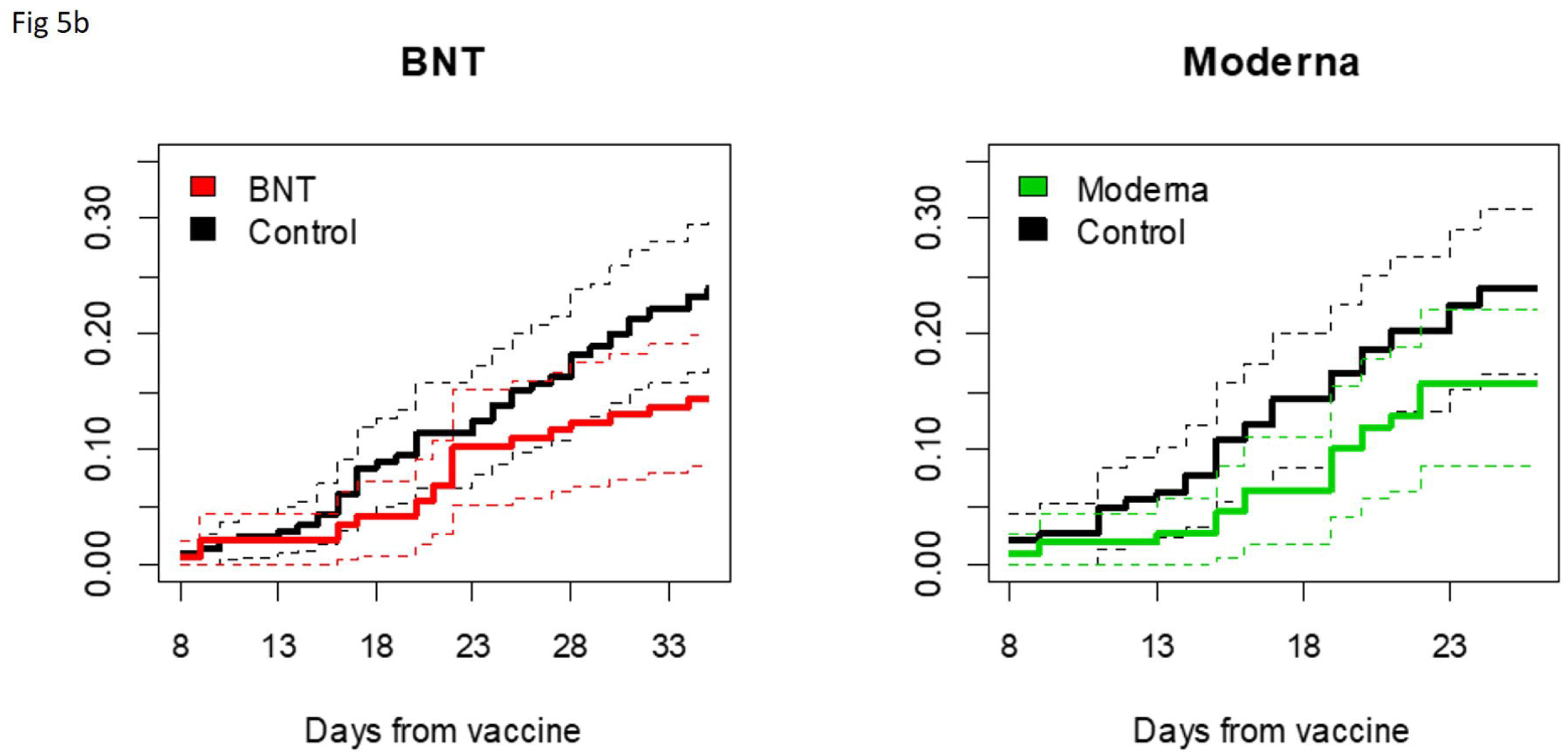
Cumulative incidence of a. all SARS-CoV-2 infections and b. symptomatic SARS-CoV-2 infections, among BNT162b2 and mRNA1273 recipients and their matched controls. 95%CI are depicted by the dotted lines.

### Cumulative Incidence and Vaccine Efficacy

In total, 29 and 28 of BNT162b2 and mRNA1273 recipients, respectively, were infected by SARS-CoV-2 during the study period; of these, 28 and 24 were defined as breakthrough infections (i.e., from day 8 after the fourth dose), and 22 and 17 were defined as breakthrough symptomatic disease. Of the 426 participants in the control group, 308 served as matched controls for BNT162b2 recipients and 239 as matched controls of mRNA1273 recipients (54 served as controls for both groups). Of the 426 controls, 181 were censored at some point, since they received a fourth vaccine dose that was approved for HCW on Jan 2, 2022. A total of 79 of the included controls were infected during the full period, but only 73 cases that occurred after day 8 of the study period (between Jan 5, 2022, or Jan 13, 2022, for BNT162b2 and mRNA1273 respectively, until Jan 30, 2022) were included in the analysis. Of all breakthrough infections, 19 were totally asymptomatic during the 7-day follow-up since their first positive SARS-CoV-2 result. In the majority of cases (65-72% in both groups) symptoms were mild (without fever of ≥38°C). Reports of fever that lasted less than 48hr was uncommon, and no fourth dose vaccine recipients reported fever that lasted for >48h. However, 19% of the BNT162b2 control group and 9% of the mRNA1273 control group reported fever that lasted >48 hours (Table 1). While symptoms, in all groups, were mostly mild to negligible, the vast majority had a relatively high viral load (with GMT of N-gene Ct of 25.3 (95%CI: 22.8-28.1), 25.1 (95%CI: 22.1-28.5) and 24.8 (23-26) in the BNT162b2, mRNA1273 and control groups, respectively) and thus, breakthrough cases were presumably infective. (**Table 1)**

**Table 1:**
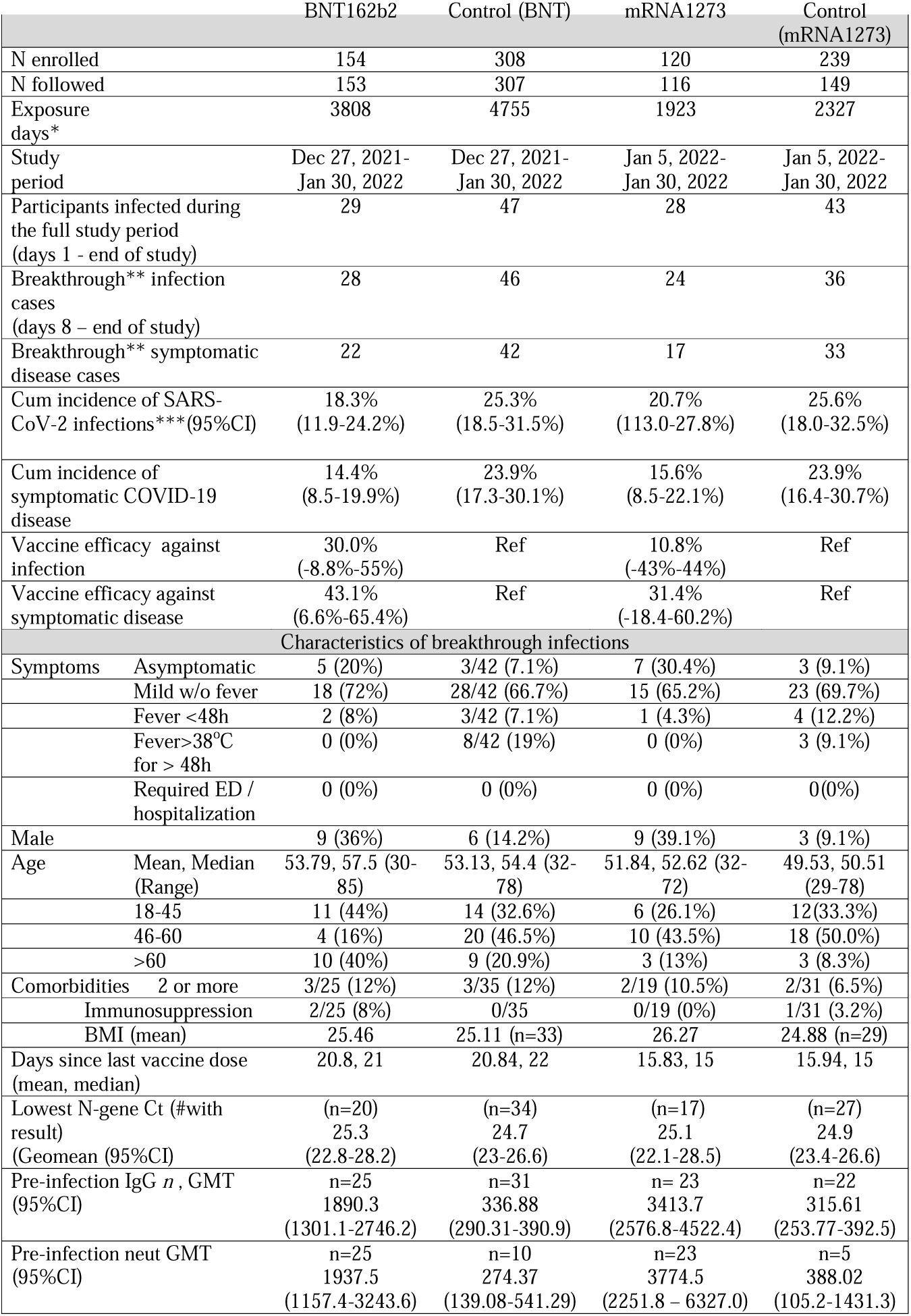
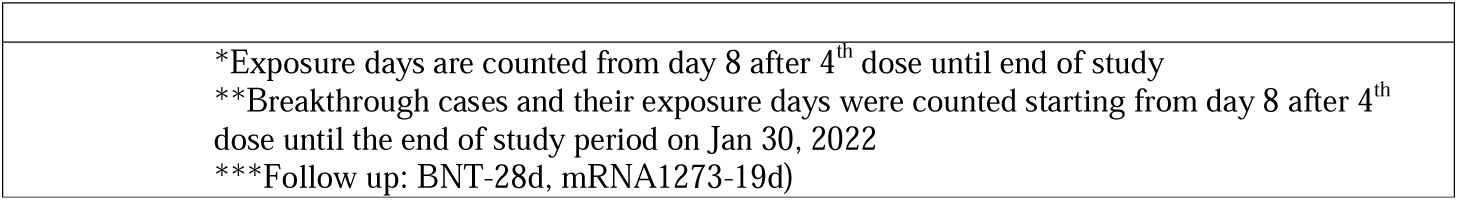

The cumulative incidence over the period from 8 days after receipt of BNT162b2 until the end of study (maximum follow up of 29 days) was 18.3% (95% CI: 11.9 to 24.2%), compared to 25.3% (95% CI: 18.5 to 31.5%) among their controls. The cumulative incidence over the period from day 8 to 23 days after receipt of the mRNA1273 vaccine was 20.7% (95% CI: 11.3 to 27.8%), compared to 25.6% (95% CI: 18.0 to 32.5%) among the mRNA1273 controls. (Note that the period of exposure for those receiving mRNA1273 was later and with a higher background incidence than for those receiving BNT162b2, so the cumulative incidences for these two vaccine groups cannot be compared.) With adjustment for period of exposure and age-group, for all SARS-Cov-2 infections, the vaccine efficacy was 30% (95% CI: -9 to 55%) for BNT162b2 and 11% (95% CI: -43 to 44%) for mRNA1273 (Table 1 & Figure 4a and 4b). For symptomatic disease, the vaccine efficacies were 43% (95% CI: 7 to 65%) and 31% (95% CI: -18 to 60%), respectively.

## DISCUSSION

This open-label, clinical trial was designed to assess the immunogenicity and safety of a fourth dose of two mRNA vaccines, BNT162b2 or mRNA1273, administered four months after the third dose in a series of three BNT162b2 doses (given on days 1, 21 and a booster dose 5-6 months later). As secondary outcomes, we also assessed the cumulative incidence of all infections as well as symptomatic disease and calculated vaccine efficacy of the fourth dose (of either vaccine) compared to three doses of BNT162b2.

The major strengths of this study include the availability of the serologic history of the participants since initial vaccination, very intensive and meticulous follow-up, which included active weekly SARS-CoV-2 PCR testing, detailed information on comorbidities, vaccine history, serology history, symptoms and adverse events.

Our study found that the fourth dose did not lead to significant adverse events despite triggering mild systemic and local symptoms in the majority of vaccine recipients. Since the fourth dose was approved in Israel for individuals over 60 years old, HCW and immunocompromised populations above 18 years, future studies will investigate the safety of the fourth dose in larger cohorts. Nevertheless, in light of numerous studies investigating the safety and reactogenicity of one, two and three mRNA vaccine doses^7,11,15–17^, our results suggest that the safety profile of the fourth dose is likely similar to that of previous doses. Interestingly, greater reactogenicity in younger adults compared to those aged 60 or more, which was previously reported, was mostly observed in the BNT162b2 recipients and less so in the mRNA1273 recipients. Yet, our study was not powered to identify less than 20% difference in AE rates.

Our study was designed primarily to determine the immunogenicity of a fourth dose and to assess whether an mRNA heterologous fourth boost (i.e., mRNA1273 following three BNT162b2 prior doses) would be more immunogenic. Our results clearly show that both mRNA vaccines significantly induce IgG and neutralizing antibodies. Moreover, both vaccines induced ∼10 folds the specific neutralizing response against Omicron and other VOC. As antibodies are found to be correlates of protection^18–20^ our serology results including the specific neutralizing ability against Omicron and the comparison to the third dose can also project on vaccine protection. Comparing the initial response to the fourth dose with the peak response following a third dose, did not demonstrate substantial differences in humoral response or in the amount of Omicron specific neutralizing antibodies (this study and Nemet et al.^15^). Overall, these data raise the possibility that the fourth dose does not boost immunity but simply restores it to peak levels. It is yet to be observed whether the waning of this fourth dose will be at a similar rate as that observed after the third dose and whether it will differ between the two mRNA vaccine groups.

While our study was not originally designed to assess vaccine efficacy, which was only a secondary outcome, the rapid spread of Omicron and the meticulous study design, with rigorous active surveillance of all infections, allowed us to determine cumulative incidence following a fourth dose and assess vaccine efficacy despite the relatively small cohorts used. Overall, during the study period 25% of the control groups were infected by SARS-CoV-2 and 18-20% of the vaccinated groups had concurrent breakthrough infections, leading to a low vaccine efficacy against infections of 11-30%, with relatively wide confidence intervals. Moreover, most of the infected HCW, in all groups complained of only negligible symptoms, which in many cases would not have been tested or reported, without the active surveillance. Yet, most of these infected HCW were potentially infectious, with relatively high viral loads. Thus, the major objective for vaccinating HCW was not achieved. The increased efficacy against symptomatic compared to asymptomatic infections found in this study suggest that the fourth dose may be more efficacious against severe disease and death, as was recently observed^22^. Therefore, older and vulnerable populations who are at higher risk for severe disease may benefit most from a fourth vaccine dose.

Our study has several limitations. First, this was not a randomized placebo-controlled trial, since it was primarily designed to assess immunogenicity, which should not be affected by our study design. However, generating potential biases in assessing vaccine efficacy. To overcome these, we compared each intervention group separately to an age-matched 2:1 control group. Second, the two intervention arms were initiated with a one week difference, leading to two potential biases; time dependent, due to the rapid surge during the study period, and minor difference in the population baseline characteristics of each arm (e.g., slightly younger in the mRNA1273 cohort). These potential biases were addressed by using a Poisson model accounting for calendar time and age. While we did not find a significant difference between the two mRNA vaccines, this is an interim report, and differences in durability of the vaccine effects may be identified only with future follow up. Finally, our study is relatively small, and thus wide confidence intervals for vaccine efficacy are reported.

Our data provides evidence that an mRNA fourth vaccine dose is immunogenic, safe and somewhat efficacious, apparently more against symptomatic disease. Four to five months after the third dose, the fourth dose increases immunogenicity and restores it to levels comparable to peak antibody levels after the third vaccine dose. Thus, while mRNA vaccines seem to be highly potent and protective against severe disease, next-generation vaccines may be needed to provide better protection against infection with highly transmissible future variants. Continued monitoring of the immune response will allow to assess the durability of the two vaccines and identify which population may benefit from it.

## Data Availability

All data produced in the present study will be available upon reasonable request to the authors

## Acknowledments

We thank the Sheba HCW participating in the trial for their full cooperating. We thank the members of the safety monitoring committee, led by Naty Keller. We thank the Sheba management, particularly Dr. Amir Grinberg, Anat Peles, Limor Ben-David, Bella Ben Mordechai and their team for their assistance. Vered Roa, Efrat Steinberg and Yael Beker-Ilani for coordinating the study, recruitment and follow up. We thank the laboratory technicians for the laborious work, Ravit Koren,Tal Levin, Osnat Halpern, Yara Kanaaneh, Shiri Kats-Likvornik, Alex Aydenzon, Hanaa Jaber, Lilac Or and Mayan Atias.

